# Patients’ Perspectives on the Contributors to and Impact of Delayed ACL Injury Diagnosis: A Qualitative Study

**DOI:** 10.64898/2026.06.23.26355683

**Authors:** Martin Thomas, Colin Ayre, Nick Dobbin, Tom Hughes

**Affiliations:** Harrogate and District NHS Foundation Trust, Harrogate, UK; School of Allied Health Professions and Sport, The University of Bradford, Bradford, UK; Bradford Teaching Hospitals NHS Foundation Trust, Bradford, UK; Department of Health Professions, Faculty of Health and Education, Manchester Metropolitan University, Manchester, UK; Institute of Sport, Manchester Metropolitan University, Manchester, UK

**Keywords:** Healthcare access, Diagnostic pathways, Patient experiences, Healthcare communication, Patient centred care

## Abstract

Anterior cruciate ligament (ACL) injuries are associated with physical, psychological and social consequences, yet diagnosis is often delayed. Existing research has focused on clinical and organisational contributors to delayed diagnosis, with limited attention on patients’ lived experiences of the diagnostic process. This study aimed to explore patients’ perspectives on the contributors to and impact of delayed ACL injury diagnosis. A qualitative study was conducted, informed by a critical realist perspective, using semi-structured interviews with ten UK-based adults who experienced a diagnostic delay exceeding three months following ACL injury. Data were analysed using codebook thematic analysis. Three interrelated themes were generated: understanding and interpreting the injury experience; navigating healthcare pathways and professional interactions; and the impact of delay, recovery and life disruption. Participants described early uncertainty, symptom normalisation and attempts to self-manage the injury, often influenced by competing work, study or family commitments. Delayed diagnosis was shaped by fragmented healthcare pathways, inconsistent advice, repeated consultations without progression and perceived dismissal of patient concerns. MRI and specialist consultations were commonly viewed as pivotal moments that validated the injury and diagnosis. Delayed diagnosis had substantial consequences extending beyond physical symptoms, including disrupted sport participation, altered occupational and parenting roles, psychological distress, and a perceived loss of time and opportunity. Diagnostic delay following ACL injury appears to arise through the interaction of patient decision making, clinical encounters and healthcare system constraints. Improving timely diagnosis may therefore require person-centred approaches that strengthen public awareness, support clearer communication and improve continuity and access within healthcare pathways.

## Introduction

Anterior cruciate ligament (ACL) injuries are a growing global health concern, with incidence rising internationally. (Bram et al., 2021) Current incidence rates are estimated to be 69 to 78 per 100,000 person years, (Nordenvall et al., 2012; Sanders et al., 2016) equating to approximately 690 to 7,800 new ACL injuries annually in small countries (1 to 10 million population), rising to approximately 34,500 to 390,000 in large countries (50 to 500 million population). Management of ACL injuries commonly includes exercise rehabilitation or surgical ACL reconstruction (ACLR) combined with rehabilitation, both of which can improve knee function, activity participation, and quality of life, while reducing reinjury risk. (Filbay & Grindem, 2019) Early and accurate diagnosis is therefore critical to enable timely intervention.

ACL injuries typically present with key clinical features at the time of injury that can facilitate early diagnosis, including a giving way episode, a perceived or audible pop or crack, immediate inability to continue activity, and rapid joint effusion. (Ayre et al., 2017) However, despite advances in clinical assessment and imaging, diagnosis remains challenging, particularly in the acute injury phase. (Clifford et al., 2021; Guillodo et al., 2008) Historically, diagnostic delays of up to 22 months have been reported, (Bollen & Scott, 1996) although more recent evidence suggests improvements have been made, with mean delays of 1.5 months. (Allott et al., 2022) However, up to 22% of patients still experience delays exceeding one year. (Parwaiz et al., 2016) Diagnostic delay may typically reflect healthcare system factors rather than delayed presentation, as most patients (52-81%) seek healthcare within days of injury, mainly via emergency departments (EDs). (Clifford et al., 2021; Perera et al., 2013) Acute clinical examination is often pain limited, which reduces diagnostic accuracy, (Whittaker et al., 2020) and key clinical features may be misinterpreted by practitioners with limited specialist musculoskeletal training or expertise. (Allott et al., 2022) Recent evidence suggested that patients with an ACL injury also experienced substantial variation in the UK’s National Health Service (NHS) diagnostic pathways, ranging from rapid referral to specialist services from EDs, to being advised to self-manage for prolonged periods before seeking further GP advice. (Carter et al., 2024) Qualitative studies across a range of knee injuries found that patients similarly perceive diagnostic delay to be attributed to organisational shortcomings, inefficient referral processes and poor communication across healthcare settings, (Holm et al., 2023; Watkins et al., 2020) often resulting in frustration. (Holm et al., 2023; Scott et al., 2018; Watkins et al., 2020)

Delayed ACL injury diagnosis may have important consequences for patients, healthcare systems and society. Although the independent effect of diagnostic delay on long-term outcomes remains unclear, early diagnosis enables timely intervention, which can target modifiable risk factors for post-traumatic osteoarthritis (e.g., physical activity and quadriceps strength) and reduce reinjury risk. (Filbay & Grindem, 2019) This is important, because recurrent instability and reinjury are associated with increased risk of secondary meniscal and chondral injury, (Arastu et al., 2015; Snoeker et al., 2013) which in turn are linked to an elevated risk of post-traumatic osteoarthritis (21-48%) compared with isolated ACL injury (0-13%). (Lie et al., 2019) These physical consequences are reflected in patient outcomes, with around half of patients not returning to sport (Filbay et al., 2025) and a third reporting persistent limitations affecting employment at two years. (von Essen et al., 2020) Quality of life (QoL) is frequently reduced, (Ardern et al., 2014) especially prior to receiving definitive treatment, (Larose et al., 2022) contributing to a considerable socioeconomic burden. (Filbay et al., 2022)

Beyond physical consequences, ACL injury and management can have detrimental effects on emotional and social wellbeing, as well as athletic identity. (Kaplan et al., 2026) Psychological effects such as fear of re-injury, anxiety, and depression are common, (Ardern et al., 2016; Ardern et al., 2014; Filbay & Grindem, 2019) which may negatively influence rehabilitation engagement and return-to-sport outcomes. (Ardern et al., 2016) Existing evidence also suggests that delays early in the care pathway may be an important contributor to further anxiety and distress. (Robling et al., 2009)

Understanding patient experiences during this diagnostic phase is therefore essential, as timely diagnosis, patient-clinician interactions and organisational processes can all meaningfully influence patient outcomes. Understanding where and why delays occur may help identify points in the pathway where improvements could be most effective. However, despite this, existing research has largely focused on patients who have undergone ACL reconstruction (ACLR) and their experiences of their broader injury management journey. (Kaplan et al., 2026) Only one qualitative study has examined patient experience during the pre-surgical phase, conducted in patients where ACL diagnosis had already been made. (Robling et al., 2009) Thus, early patient experiences following injury, particularly during initial healthcare interactions and diagnostic pathways, remain underexplored. (Kaplan et al., 2026) This is especially important for individuals experiencing prolonged diagnostic delay (>3 months), whose needs and experiences are poorly understood.

This study aims to explore the lived experiences of UK-based patients who experienced a diagnostic delay exceeding three months following ACL injury, examining (1) experiences, responses, attitudes and perceived contributors to diagnostic delay, and (2) the perceived impact on physical, psychological, and social well-being. The findings may inform changes in diagnostic processes and support the development of more efficient and timelier patient-centred ACL injury management pathways.

## Materials and Methods

### Study Design

This qualitative phenomenological study explored patients’ experiences of diagnostic delay following anterior cruciate ligament (ACL) injury. Reporting was in accordance with the Consolidated Criteria for Reporting Qualitative Research (COREQ; Supplement 1). (Tong et al., 2007)

The study was informed by a critical realist perspective, which conceptualises reality as three domains: the empirical (patients’ experiences and observations), the actual (events occurring regardless of observation), and the real (underlying mechanisms that generate these events). (Stutchbury, 2022) This perspective informed the analysis by exploring both patients’ experiences of delayed ACL injury diagnosis and the healthcare processes and mechanisms contributing to diagnostic delay.

Ethical approval was obtained from the London Riverside Research Ethics Committee (Ref: 17/LO/1489). All participants provided written informed consent.

### Researcher Positioning and Reflexivity

All interviews were conducted by the first author (MT), a specialist musculoskeletal NHS physiotherapist with six years’ experience managing ACL injuries and interest in this area. The project was submitted as part of an MSc study programme at the University of Bradford, supervised by CA with 25 years’ experience of managing ACL injuries. MT and CA’s clinical backgrounds informed the development of the research question and interview schedule. TH provided clinical expertise in knee injury diagnosis, with 23 years’ experience. ND, who has experience in qualitative research but no clinical background in managing ACL injuries, acted as ‘critical friend’ and reviewed the transcripts, coding process, and construction of themes. All authors were male.

Reflexivity was incorporated throughout the research process. Field notes were recorded by MT following each interview to document contextual observations and early analytic reflections. Reflexive memos were also maintained during coding and theme construction. This process supported transparency in the development of themes and encouraged ongoing reflection on how researcher knowledge and assumptions may have influenced analysis.

### Participants and Recruitment

Purposive sampling was used to recruit adults (aged >18 years) with confirmed ACL injury (verified by magnetic resonance imaging, arthroscopy or diagnosis by a specialist clinician) who experienced delays of at least three months between injury and diagnosis. No upper limit for delay duration was imposed to capture a range of experiences, including prolonged diagnostic uncertainty. Individuals with concomitant injuries (e.g., multiple ligament injuries, neurovascular injuries), unclear injury mechanisms and those treated by the primary researcher were excluded.

Participants were recruited from three National Health Service sites: Croydon University Hospital, Bradford Teaching Hospitals Trust and Airedale Hospitals Trust. Eligible individuals attending physiotherapy or orthopaedic clinics were identified and approached by a member of the clinical team and provided with study information. Individuals who expressed interest were contacted by the lead researcher (MT) to arrange interviews and provide written consent. Recruitment continued until sufficient data richness was achieved to address the research aims.

### Data Collection

Data were collected between February and September 2018. Data were generated through semi-structured interviews. The interview guide (Supplement 2), informed by relevant literature and study aims, explored three broad areas: (1) the initial injury event and interpretation of symptoms; (2) experiences of seeking healthcare and interactions with healthcare professionals; and (3) the impact of delayed diagnosis on physical, psychological and social domains. Open-ended questions encouraged participants to describe their experiences in their own words, with prompts used to explore issues in greater depth. Participants were also asked to reflect on factors they believed contributed to delay and how the diagnostic pathway could be improved. The interview guide was piloted between the lead researcher and project supervisor.

Single interviews were conducted in-person, with only the participant and interviewer present, typically alongside participants’ clinic appointments in a private clinic room. Conducting interviews within the clinical environment minimised travel and scheduling burden while providing a familiar, quiet setting conducive to discussing healthcare experiences. (McGrath et al., 2019) However, the clinical setting may also have influenced interview dynamics and participants’ willingness to express critical views as medical environments can reinforce perceived power differentials that may affect openness when discussing sensitive issues. Although the interviewer was not involved in participants’ clinical care, the healthcare setting may still have shaped perceptions of authority and influenced participants’ accounts.

### Data analysis

All interviews were audio recorded, transcribed verbatim, and anonymised prior to analysis. Data were analysed using codebook thematic analysis as described by Braun and Clarke. (Braun & Clarke, 2021) Analysis began with familiarisation through repeated reading of transcripts and field notes. Initial coding identified segments of text relevant to the research aims using NVIVO (Lumivero, Version 14, 2023). A preliminary coding framework was developed using sensitising concepts from the interview guide and literature alongside inductively generated codes emerging from the data. These codes were organised into a codebook that included code definitions and illustrative examples. Codes were compared across transcripts to identify patterns of meaning. Themes were primarily derived inductively from the data, while being informed by sensitising concepts from the interview guide and existing literature. Themes were reviewed and refined through repeated engagement with the dataset. Final themes were defined and named to capture key patterns relating to diagnostic delay and its consequences.

### Trustworthiness and Rigour

Trustworthiness was considered in relation to the principles of credibility, dependability, confirmability and transferability. Credibility was supported through prolonged engagement with the data, iterative coding and use of a peer reviewer. Response validation was not used. Dependability was supported through the structured codebook that guided the coding process. Transferability was assisted through detailed description of the research context, participant characteristics and use of detailed and rich quotes. Confirmability was enhanced through reflexive practices and through grounding themes in illustrative participant quotations.

## Results

Ten participants (six male, four female) participated in the study, which reflected all of those invited, with interviews lasting 15 to 35 minutes. Participants were aged between 18 to 35 years at the time of interview and identified as White British, Irish, Eastern European and Asian. Participants described a range of injury circumstances and healthcare pathways across three National Health Service settings and, in some cases, private healthcare services. The self-reported duration of diagnostic delay varied between 3 months to 10 years and reflected differences in injury presentation, access to healthcare services, and clinical decision making within the diagnostic pathway. Four of the participants had diagnostic delays of more than a year.

Analysis generated three interrelated themes that describe how participants interpreted their injury, navigated healthcare pathways and experienced the wider consequences of delayed diagnosis. The themes reflect the progression of the injury experience from the moment of injury, through encounters with healthcare professionals and healthcare systems, to the longer-term physical, psychological and social impact of prolonged instability and delayed treatment (Table 1).

**Table 1.** Overview of the codebook structure.

| Theme | Key Codes |
| --- | --- |
| Understanding and interpreting the injury experience | • Immediate injury interpretations<br>• Audible or sensory indicators of injury<br>• Self-management and continuation of activity<br>• Early uncertainty and downplaying injury severity<br>• Feeling dismissed or not believed by clinicians |
| Navigating healthcare pathways and professional interactions | • Fragmented healthcare pathways<br>• Repeated assessments without progress<br>• Inconsistent clinical advice<br>• Delays in imaging or referral<br>• Positive encounters and effective clinical decision making |
| The ongoing impact of delay, recovery and life disruption | • Loss of sport participation<br>• Occupational disruption<br>• Parenting and family impact<br>• Psychological consequences of delayed diagnosis<br>• Perceived loss of time and opportunity |

## Theme 1: Understanding and interpreting the injury experience

This theme explores how participants made sense of their injury following the initial event. Participants described drawing on bodily sensations, previous experiences of injury and the reactions of others to interpret what had happened. These early interpretations shaped decisions about whether medical care was required and influenced how individuals subsequently engaged with healthcare services. Participants frequently described attempts to maintain everyday activities despite ongoing symptoms, particularly when the seriousness of the injury remained uncertain.

### Immediate interpretations and certainty

For many participants, the injury event was experienced as clearly abnormal, with accounts describing intense pain, instability or unusual sensations that suggested something significant had occurred. These interpretations were grounded in embodied experiences rather than formal clinical knowledge. Several participants described pain as the defining feature of the injury, often unlike anything previously experienced. For example, P1 explained that ‘I had never experienced that sort of level of pain before. I thought it was broken completely’.

Others identified instability as the key indicator, describing repeated episodes of the knee giving way following a twisting mechanism. Similarly, some participants reported hearing or feeling a distinct, audible crack or pop at the moment of injury, reinforcing perceptions that the joint had been seriously damaged. For example, P7 recalled hearing ‘an almighty crack and pop’, while P4 reported that ‘my knee popped and everybody heard it’. In both cases the sensation was followed by rapid swelling and an immediate perception that the knee had moved abnormally. Indeed, P3 explained that when she attempted to step forward the leg seemed to disappear beneath her, while P8 recognised the sensation immediately due to a previous ligament injury in the opposite knee.

### Early uncertainty, downplaying severity self-management and patient-initiated delay

Despite initial concerns, many participants attempted to normalise or minimise the severity of the injury in the days that followed. Several described expecting symptoms to resolve spontaneously and managing the injury independently, often drawing on prior experience. For example, P6 drew on a previous injury to guide self-management:

> I had physio for my ankle before and I just kind of, I know how to…the kind of…like give myself a kind of physio thing. I just thought I would ease myself into using my leg again. So, after like 8 months I played again, I played football again, of course after…initially…like half a minute I tried to change direction and my leg just went.

Practical pressures, including work and study commitments, further shaped early decision making. Some participants (e.g., P3, P7) prioritised maintaining routine activities despite ongoing symptoms, even when this conflicted with informal advice or physical limitations.

> I bought myself a stick and I thought, I felt a little bit like a fraud. I know it is not broken but if I had been told that be careful, don’t step on that leg or at least take it easy but I thought to step on that leg as much as I can and ignore the pain. (P3)

In some cases, the perceived mechanism of injury contributed to a downplaying of severity. P6 initially believed she had simply twisted the knee and attempted to return to activity prematurely before experiencing repeated episodes of instability. P10 similarly managed the injury as a minor sprain until persistent symptoms prompted further medical assessment. As symptoms continued, some participants began to question their own judgement.

External factors, such as pre-existing commitments, also constrained opportunities to seek further assessment, extending the period before diagnosis. For example, after attending hospital the day after the injury, P1 noted:

> They just said it might be like a sprained knee or something like that. I was quite relieved at that first as it wasn’t anything as serious as I initially thought I went out to Australia five days after that injury. So didn’t really…I didn’t have time to delve into it further with the NHS. So, it wasn’t a good situation.

### Contesting legitimacy: feeling unheard or dismissed

Several participants described clinical encounters where professional reassurance conflicted with their lived experience of symptoms, particularly when normal radiographs appeared to conclude assessment despite ongoing instability. P3 for example, presented to hospital after hearing a crack and being unable to weight-bear, but was advised to take analgesia and try to walk, leading to confusion: ‘I started doubting myself and tried to do what that man said, but then I thought it is impossible. I felt confused about what to do’. Others described feeling that their concerns were not fully acknowledged, particularly when symptoms persisted across multiple consultations:

> I don’t know if I could take any positives from it. Negatives are that there was a problem, and I knew what the problem was, as I had previous experience. It got brushed aside, you know. Didn’t listen to what I said, and it turns out that it was ACL….No… No not at all, because it happened with my right knee. Eh… I went in, they had a look and said you are alright. There is not a problem. In two weeks, you can go back to your sports and I said okay. As it happens, I give it three, four weeks, I went back and I did more damage than what originally was done. No, I didn’t feel… I didn’t feel confident with the advice. (P8)

When asked about how this made them feel, they went on to state:

> I don’t think that there was anything that left me satisfied with. The only thing I could say that, is that I was seen by somebody in the NHS. What left me unsatisfied was that I had known what I had done, it was not getting treated well… as well as it should have been. How many professionals?… a handful maybe three or four and then, I went in for an MRI and that’s what diagnosed it. Because every consultation I had with the professional, they do the tests with the knee, check all the ligaments. For some reason, they could not find that my ligaments were weak. They check all the ligaments of my knee and they will be like… no your ligaments are fine. Only knew it when I did the MRI. (P8)

In contrast, some participants described later encounters with specialists as more definitive, highlighting variation in clinical assessment across the pathway:

> But you think that a physio would be more experienced in examining a knee, a sports physio, than a GP. So, the only other thing is that it is a surprise that 2 people didn’t pick it up, but when I was examined by the surgeon, I could feel it, I could see it. I could almost remember the words he said “there was no question that your cruciate is gone” in a very sort of blunt manner. I say, “do we need a scan” he said, “no we don’t need a scan it is definitely gone” and I could appreciate that it is gone. I was thinking to myself why could this not… why could someone not elicit that same examination finding. Because I know people had done the same anterior drawer, repeatedly over the previous 3 months, nobody had managed to make a clunk like that guy could, so it was literally a 5 mins, if even consultation, you know. So, yeah guess the main thing is a little bit of bad luck that for whatever reason the 2 people I saw didn’t manage to pick it up, maybe I was more tense I don’t know. (P9)

## Theme 2: Navigating healthcare pathways and professional interactions

Participants’ experiences of navigating healthcare pathways were often characterised by complexity, repetition and uncertainty. Rather than a clear and linear diagnostic process, many described being passed between practitioners, receiving inconsistent advice and experiencing delays in accessing imaging or specialist care. These experiences shaped participants’ understanding of their injury over time and influenced their confidence in clinical decision making.

### Navigating care: challenges, support and key investigations

A consistent feature across accounts was a fragmented pathway, characterised by repeated consultations without progression towards diagnosis. P1 described being passed between practitioners without continuity:

> I was obviously not sure how many different levels of consultants you have to see but I was just being passed on from one person to the next person, to the next person, to the next person… like a doll. You shouldn’t need 8, 9, 10 people to tell me that I have the same injury before I have it done.

Participants also highlighted the need for persistence in accessing appropriate investigation, particularly magnetic resonance imaging (MRI).

> I feel that the GP appointment was not very well informed in terms of what is wrong with my knee and what potentially could be wrong with it and how. The GP from that main episode that happened, I said to him “Why are you sending me for an x-ray?” It is not going to bone. I have not broken a bone. It is a ligament issue; you need to send me for an MRI scan”. P2 went on to the suggest that “For two years I kept asking for an MRI… I felt neglected. (P2).

For many participants, MRI represented a critical moment that validated concerns and provided diagnostic clarity. P1 described this shift: ‘As soon as that MRI scan is done… that is the confirmation of the injury. You can argue with someone pulling on your knee, but you cannot argue with an MRI.’

While many participants described delays, others experienced more efficient and coordinated care pathways characterised by timely referral, clear communication, and continuity across services. These accounts highlighted early clinical recognition, active prioritisation within the system, and collaborative discussions around management options once specialist care was reached. For example, one participant described a seamless progression from emergency department attendance to physiotherapy support:

> I think probably, just… my most recent experience has been so overwhelmingly positive in that, it was just… even with the really long wait times in A&E, I was seen quickly, I was X rayed. Somebody consulted my case, I got the same treatment I had the first two times, but then it was followed up with a physio appointment… I think it was a week later, so seven days. (P5)

### Inconsistent advice and clinical uncertainty

Participants frequently described experiences of receiving unclear or conflicting guidance about activity and recovery following their injury. Several reported that advice and communication was either vague or inconsistent across consultations, creating confusion about which activities were safe or not. For some, the absence of clear explanations meant they were left to make decisions on their own, sometimes resulting in actions that they perceived as inadvertently worsening their condition. Others described the frustration of receiving differing opinions from multiple practitioners, which undermined their confidence in the guidance and made it difficult to interpret the seriousness of their injury. One participant captured the challenge of navigating unclear advice:

> I was told no football but not what movements to avoid… so in my head I thought as long as I am not playing, I am okay. But then I was still doing other things, turning, twisting, and that is when it gave way again. I think that is where I did more damage because I just did not really understand what I should or should not be doing. (P1)

Another highlighted the confusion caused by conflicting professional opinions:

> Different people kept telling me different things… one would say it is fine, another would say just rest it, another would say try and get back to normal. You don’t really know who to believe, so you just kind of guess what to do and hope for the best. (P7)

## Theme 3: The ongoing impact of delay, recovery and life disruption

Delayed diagnosis and prolonged instability had consequences that extended beyond the initial injury. Participants described sustained disruption to sport participation, occupational roles, family life and psychological wellbeing. These impacts accumulated over time and were often intensified by the uncertainty surrounding diagnosis. Participants’ accounts highlight how delayed diagnosis was experienced not only as a clinical issue, but as a disruption to everyday life and future trajectories.

### Disruption everyday life and the experience of lost time

Delayed diagnosis and prolonged instability caused substantial disruption to participants’ everyday lives, affecting sport participation, occupational roles, and family responsibilities. For many participants, sport was central to their identity, and its loss carried immediate and longer-term consequences. P1 described withdrawing socially following their injury: ‘my entire life revolves around sport… I did not speak to anyone for a week. I just kind of shut myself away because I did not really know what to do with myself without it’. For others, this absence extended over prolonged periods: ‘I have missed ten years of netball… and it is not just the playing; it is the social side of it as well’, illustrating how disruption extended beyond physical participation into social connection and belonging. (P5)

Disruptions also reshaped occupational roles and identities. P4 described being unable to fulfil the physical aspects of their role: ‘I went from being out on the streets to being behind a desk… I hated it. It was not what I joined for’.

Family life was significantly affected where participants could not engage in everyday activities. P2 reflected on missed experiences with their children: ‘you cannot bring the time back… I have missed experiences with my kids, simple things like running around with them or even just being active with them’. P3 described how the injury affected parenting roles:

> Literally I couldn’t walk. I have a young* daughter so I couldn’t play with her. I remember after 3 weeks I would go to work and back and lay on sofa, can’t do anything… I remember my daughter standing there and crying saying, ‘I want my mummy’. I was in front of her and I said, ‘I am here’. What she meant was she wanted her old mummy who runs and plays with her… ‘Mummy can you pick me up?’ ‘No, I can’t because of my leg’. ‘Can’t we run, can’t we play?’ and I couldn’t do it. (P3)
>
> *Note that specific age has been removed and replaced with the word ‘young’.

Participants often framed their experiences in terms of lost time, linking delays in diagnosis to missed opportunities and altered life trajectories. P1 reflected on the impact during a key developmental period: ‘Learning I would not play properly until 21… it felt like losing formative years’. P7 described delays affecting sport and education: ‘It took so long to actually get the diagnosis and then even longer to get anything done about it… it just felt like everything was on hold for so long.’ Broader lifestyle changes were also evident. P3 reflected: ‘I had a very active lifestyle… but after work I couldn’t do anything. My lifestyle changed, I put weight on… from being very active I was just sat in one place.’

### Psychological consequences and loss of confidence

Participants described a substantial psychological impact associated with both the injury and delays in diagnosis, often linked to uncertainty and an inability to return to normal activities. For example, P8 described how this affected both mental wellbeing and daily life:

> It made me feel down to be honest, because I was not able to do anything. No gym, no football, no walk… just nothing. It just felt like everything had kind of stopped. You just feel stuck… like you cannot move forward with anything because you do not actually know what is wrong properly. (P8)

Alongside this, repeated instability contributed to a reduced sense of trust in their knee. P9 described how ‘every step you take you are wondering if it is going to go again’, while P6 explained that she was ‘always second guessing’ any movement due to risk.

## Summary of Results

These findings illustrate how delayed diagnosis of ACL injury unfolds across individual, clinical and structural levels. Early interpretations of injury, shaped by embodied sensations and prior experience, influenced initial help seeking behaviours and expectations of recovery. These interpretations were subsequently negotiated within healthcare encounters, where inconsistent advice, fragmented care pathways and limited access to imaging contributed to prolonged uncertainty and, in some cases, a perceived need for patients to advocate for further investigation.

Across the dataset, diagnosis was not experienced as a single clinical event but as the outcome of a process, shaped by repeated interactions with healthcare professionals and systems. In this context, MRI and specialist consultation often represented pivotal moments that validated participants’ experiences and provided diagnostic clarity. However, by the point of confirmation, many participants had already experienced extended periods of instability, uncertainty and disruption.

The consequences of delayed diagnosis extended beyond the clinical pathway, affecting participants’ physical functioning, psychological wellbeing and broader life trajectories. Disruptions to sport, work and family roles were frequently accompanied by reduced confidence, emotional distress and a sense of lost time. These impacts were often cumulative, reflecting the interaction between ongoing symptoms and delays within the healthcare system.

Taken together, the themes highlight that diagnostic delay is not solely attributable to individual clinical encounters but emerges through the interaction between patients’ perception and interpretations of the injury and its seriousness, clinical decision making and structural features of healthcare pathways, as well as patients’ external commitments. From a critical realist perspective, participants’ accounts provide insight into how these underlying mechanisms shape both the experience and consequences of delayed diagnosis in practice.

## Discussion

This study investigated patients’ experiences of diagnostic delay following ACL injury, and its physical, psychological and social consequences. Our findings suggest diagnostic delay does not arise from a single missed clinical event but is a cumulative process shaped by interactions between patient, clinical, and structural factors. While previous literature has largely attributed delayed diagnosis to organisational and structural inefficiencies (Carter et al., 2024; Holm et al., 2023; Watkins et al., 2020) or challenges in the clinical assessment of acute knee injury, (Allott et al., 2022; Parwaiz et al., 2016; Simonsen et al., 1984; Whittaker et al., 2020) our findings suggest a more complex picture, involving ongoing negotiation between patient’s lived experience of the injury and healthcare system constraints.

Across different conditions, care settings and patient populations, patient experience has been found to be associated positively with both treatment outcomes and patient safety. (Doyle et al., 2013) Although some positive experiences were reported, participants predominantly described fragmented pathways, inconsistent advice and prolonged diagnostic uncertainty, which negatively affected psychological wellbeing and perceived care quality. These findings are consistent with previous research examining ACL (Allott et al., 2022; Carter et al., 2024; Maher et al., 2025; Watkins et al., 2020) and acute knee injury pathways. (Robling et al., 2009; Watkins et al., 2020) Collectively, this evidence suggests that structural and organisational variability and insufficiencies in care pathways, remain persistent challenges for healthcare services and may contribute to regional inequities. Similar patient experiences reported in Denmark (Holm et al., 2023) also suggest that these challenges may extend beyond the UK. This reinforces the need for initiatives such as Getting it Right First Time, which aims to reduce unwarranted variation in care quality across the NHS. (National Health Service England, 2024)

Healthcare seeking behaviour is recognised as a complex process shaped by clinical, system-level, social and cultural factors. (Greenhalgh et al., 2026) Our findings extend this understanding by demonstrating how patient-level factors actively contribute to diagnostic delay following ACL injury. Symptom normalisation, competing life priorities and trust in professional reassurance emerged as key influences on delayed healthcare-seeking. These findings position patients not as passive recipients of delay, but as active participants within the diagnostic pathway, whose decisions interact with system constraints to shape outcomes. This more nuanced conceptualisation may help explain why 19-48 % of ACL injured patients take longer than a week to access definitive healthcare. (Ayre et al., 2017; Clifford et al., 2021; Perera et al., 2013) Further, participants in our study described persistent diagnostic uncertainty, with profound psychological and social consequences. This uncertainty disrupted family roles, life trajectories, employment and sports participation, contributing to a sustained sense of identity loss. While similar findings have been reported internationally across knee injury pathways, (Holm et al., 2023; Kaplan et al., 2026; Scott et al., 2018; Watkins et al., 2020) our results suggest that identity disruption may emerge rapidly, driven by the disabling effects of the acute injury, compounded by early diagnostic uncertainty.

Even among participants who accessed care promptly, uncertainty persisted when they felt unheard, were dismissed, or received inconsistent diagnostic explanations. Trust in physical examination findings was limited, which is unsurprising given the reduced diagnostic accuracy of ACL-specific tests in the acute post-injury phase. (Simonsen et al., 1984) These tests are often painful, (Whittaker et al., 2020) and findings may be misinterpreted when clinicians lack specialist training, as is often the case in acute care settings. (Allott et al., 2022) In contrast, MRI results and specialist consultations were perceived as pivotal events for injury validation. Collectively, these findings raise questions of the clinical utility and patient acceptability of existing early assessment practices and suggest that clinical encounters may inadvertently contribute to ongoing diagnostic uncertainty and delay.

Overall, these findings suggest that diagnostic delay following ACL injury cannot be attributed solely to healthcare system inefficiencies. Rather, delay emerges through interactions between patient decision making, clinical encounters and organisational structures. This broader conceptualisation highlights the need to consider both healthcare delivery and patient experience when seeking to improve the timeliness of ACL diagnosis.

## Implications for practice

Our findings suggest that reducing diagnostic delay following ACL injury requires approaches that extend beyond structural and clinical solutions to incorporate person□centred care, recognising patients’ lived experiences, expectations and decision□making processes. Strengthening public-facing health messaging may help discourage symptom normalisation and support earlier healthcare seeking by emphasising key injury features. Although no participants referenced NHS guidance, current recommendations advise seeking care for symptoms such as severe pain, swelling, instability, or functional limitation following knee injury. (National Health Service, 2026) However guidance does not explicitly refer to the audible or perceptible “crack” or “pop” reported in up to 74% of ACL injuries, (Ayre et al., 2017; Lukas et al., 2022) despite this being a salient feature in participant accounts.

Early clinical encounters provide an opportunity to validate patient concerns, communicate diagnostic uncertainty and offer clear safety netting advice when diagnosis is not immediately possible. From a systems perspective, improving pathway continuity and access to specialist review may reduce uncertainty and delay. Acute knee clinics may expedite diagnosis (Clifford et al., 2021; Maher et al., 2025; Parwaiz et al., 2016) although access is inconsistent and resource intensive, (Clifford et al., 2021) potentially exacerbating inequalities.

Alongside clinician education, evidence of patient□initiated delay highlights the need for simple decision support tools. However, existing tools, including the LIMP index (Ayre et al., 2017) and the Anterior Cruciate Ligament Injury Score (ACLIS), (Lukas et al., 2022) currently lack sufficient external validation for routine use.

## Strengths and limitations

A key strength of this study is its critical realist perspective, which considers the participant’s lived experience (the empirical) but also the systemic structures (the reality) that contribute to diagnostic delay. The use of semi-structured interviews supported an inductive approach, enabling themes to be constructed from participants’ accounts, while recruitment across multiple NHS settings and regions enhanced the transferability of findings.

Our study does have some limitations. The sample reflects only three geographical regions, which means care is needed when transferring these experiences to the broader ACL population within the UK and beyond. Interviews were conducted within clinical settings, which may have influenced participants’ openness due to perceived power dynamics, and the retrospective nature of data collection introduces potential recall bias.

## Future Research

Future qualitative and mixed-methods studies using larger, multi-centre samples across public and private care settings are needed to better understand variation in diagnostic experiences. Comparing patients who receive early versus delayed diagnosis may also help identify indicators of best practice. In addition, incorporating clinician perspectives could elucidate challenges to acute assessment and identify alternative approaches to early knee injury evaluation.

## Conclusion

Diagnostic delay following ACL injury appears to be a multifactorial and cumulative process shaped by interactions between patient decision making, clinical encounters and healthcare system constraints. Participants described fragmented care pathways, persistent uncertainty and inconsistent communication, which negatively affected satisfaction with the care received. Importantly, delays were not solely attributable to organisational inefficiencies, but also reflected symptom normalisation, competing life priorities and reassurance received during early healthcare encounters. These findings highlight the need for more person-centred approaches that improve early recognition, validate patient concerns and enhance pathway continuity. Addressing diagnostic delay following ACL injury will likely require coordinated improvements in public awareness, clinical assessment practices and access to timely specialist care.

## Supporting information

COREQ checklist

Interview schedule

## Acknowledgements

The authors thank all participants for their time and contribution, and the participating NHS Trusts for their support. We also acknowledge Anita Sergeant and the University of Bradford for their assistance and institutional support. We are grateful to Kamal Balakrishnan, Emma Parker, Rachel Hunt and Benoy Mathew (Croydon University Hospital), and David Tweed (Airedale General Hospital) for their contribution to this study.

## Declaration of Generative AI Use

The authors report generative AI was not used in their research or preparation of this manuscript.

## Key statements

### Data Availability

Data supporting the findings of this study are available within the research paper, presented as relevant quotations.

## Patient and Public Involvement

Patients and the public were not involved in the design, conduct, reporting, or dissemination of this study, beyond piloting the questionnaire with an individual who met the study’s inclusion criteria.

## CRediT Author Statement and ICMJE Criteria

Conceptualisation was undertaken by MT, TH, and CA. Methodology was developed by MT, CA, and ND. Formal analysis was conducted by MT and ND. Investigation was carried out by MT and CA. Data curation was undertaken by MT. The original draft was prepared by MT, TH, and ND. Review and editing of the manuscript were undertaken by all authors. Supervision was provided by CA. Project administration was undertaken by MT and CA.

All authors meet the ICMJE criteria for authorship and have approved the final version of the manuscript.

## References

Allott, N. E. H., Banger, M. S., & McGregor, A. H. (2022). Evaluating the diagnostic pathway for acute ACL injuries in trauma centres: a systematic review. BMC Musculoskeletal Disorders, 23(1), 649. 10.1186/s12891-022-05595-0

Arastu, M. H., Grange, S., & Twyman, R. (2015). Prevalence and consequences of delayed diagnosis of anterior cruciate ligament ruptures. Knee Surgery, Sports Traumatology, Arthroscopy, 23(4), 1201–1205. 10.1007/s00167-014-2947-z

Ardern, C. L., Kvist, J., & Webster, K. E. (2016). Psychological Aspects of Anterior Cruciate Ligament Injuries. Operative Techniques in Sports Medicine, 24(1), 77–83. 10.1053/j.otsm.2015.09.006

Ardern, C. L., Taylor, N. F., Feller, J. A., & Webster, K. E. (2014). Fifty-five per cent return to competitive sport following anterior cruciate ligament reconstruction surgery: an updated systematic review and meta-analysis including aspects of physical functioning and contextual factors. British Journal of Sports Medicine, 48(21), 1543–1552. 10.1136/bjsports-2013-093398

Ayre, C., Hardy, M., Scally, A., Radcliffe, G., Venkatesh, R., Smith, J., & Guy, S. (2017). The use of history to identify anterior cruciate ligament injuries in the acute trauma setting: the ‘LIMP index’. Emergency Medicine Journal, 34(5), 302–307. 10.1136/emermed-2015-205610

Bollen, S. R., & Scott, D. J. (1996). Rupture of the anterior cruciate ligament - a quiet epidemic? Injury: International Journal of the Care of the Injured, 27(6), 407–409.

Bram, J. T., Magee, L. C., Mehta, N. N., Patel, N. M., & Ganley, T. J. (2021). Anterior Cruciate Ligament Injury Incidence in Adolescent Athletes: A Systematic Review and Meta-analysis. American Journal of Sports Medicine, 49(7), 1962–1972. 10.1177/0363546520959619

Braun, V., & Clarke, V. (2021). Thematic analysis: A practical guide. SAGE Publications Ltd.

Carter, H. M., Beard, D. J., Leighton, P., Moffatt, F., Smith, B. E., Webster, K. E., & Logan, P. (2024). ‘Going through the motions’; a rich account of the complexity of the anterior cruciate ligament reconstruction pathway, a UK qualitative study. BMJ Open, 14(9), e079468. 10.1136/bmjopen-2023-079468

Clifford, C., Ayre, C., Edwards, L., Guy, S., & Jones, A. (2021). Acute knee clinics are effective in reducing delay to diagnosis following anterior cruciate ligament injury. Knee, 30, 267–274. 10.1016/j.knee.2021.04.007

Doyle, C., Lennox, L., & Bell, D. (2013). A systematic review of evidence on the links between patient experience and clinical safety and effectiveness. BMJ Open, 3(1), e001570. 10.1136/bmjopen-2012-001570

National Health Service England. (2024). Getting It Right First Time (GIRFT). (https://gettingitrightfirsttime.co.uk/ (Accessed: 17 April 2026)

Filbay, S. R., Bullock, G., Russell, S., Brown, F., Hui, W., & Egerton, T. (2025). No Difference in Return-to-Sport Rate or Activity Level in People with Anterior Cruciate Ligament (ACL) Injury Managed with ACL Reconstruction or Rehabilitation Alone: A Systematic Review and Meta-Analysis. Sports Medicine, 55(9), 2191–2205. 10.1007/s40279-025-02268-5

Filbay, S. R., & Grindem, H. (2019). Evidence-based recommendations for the management of anterior cruciate ligament (ACL) rupture. Best Practice & Research: Clinical Rheumatology, 33(1), 33–47. 10.1016/j.berh.2019.01.018

Filbay, S. R., Skou, S. T., Bullock, G. S., Le, C. Y., Raisanen, A. M., Toomey, C., Ezzat, A. M., Hayden, A., Culvenor, A. G., Whittaker, J. L., Roos, E. M., Crossley, K. M., Juhl, C. B., & Emery, C. (2022). Long-term quality of life, work limitation, physical activity, economic cost and disease burden following ACL and meniscal injury: a systematic review and meta-analysis for the OPTIKNEE consensus. British Journal of Sports Medicine, 56(24), 1465–1474. 10.1136/bjsports-2022-105626

Greenhalgh, S., Ghorbankhani, M., & Yeowell, G. (2026). Health-seeking behaviour in adults with musculoskeletal conditions: A scoping review. Musculoskelet Sci Pract, 82, 103499. 10.1016/j.msksp.2026.103499

Guillodo, Y., Rannou, N., Dubrana, F., Lefèvre, C., & Saraux, A. (2008). Diagnosis of Anterior Cruciate Ligament Rupture in an Emergency Department. Journal of Trauma and Acute Care Surgery, 65(5), 1078–1082. 10.1097/TA.0b013e3181469b7d

Holm, P. M., Simony, C., Brydegaard, N. K., Hogsgaard, D., Thorborg, K., Moller, M., Whittaker, J. L., Roos, E. M., & Skou, S. T. (2023). An early care void: The injury experience and perceptions of treatment among knee-injured individuals and healthcare professionals - A qualitative interview study. Physical Therapy in Sport 64, 32–40. 10.1016/j.ptsp.2023.08.006

Kaplan, S., Patterson, B. E., Bruder, A. M., Ezzat, A. M., Bunzli, S., & Culvenor, A. G. (2026). ’ACL - wow, this is bad’: patients’ perspectives on their anterior cruciate ligament injury and its care - a systematic review and qualitative evidence synthesis. British Journal of Sports Medicine, 60(9), 660–671. 10.1136/bjsports-2025-109746

Larose, G., Leiter, J., Peeler, J., McRae, S., Stranges, G., Rollins, M., Davidson, M., & MacDonald, P. (2022). Quality of life during the wait for ruptured anterior cruciate ligament reconstruction: a randomized controlled trial. Canadian Journal of Surgery, 65(2), E269–E274. 10.1503/cjs.007820

Lie, M. M., Risberg, M. A., Storheim, K., Engebretsen, L., & Oiestad, B. E. (2019). What’s the rate of knee osteoarthritis 10 years after anterior cruciate ligament injury? An updated systematic review. British Journal of Sports Medicine, 53(18), 1162–1167. 10.1136/bjsports-2018-099751

Lukas, S., Putman, S., Delay, C., Blairon, A., Chazard, E., & Letartre, R. (2022). Knee Ligament Sprains: Diagnosing Anterior Cruciate Ligament Injuries by Patient Interview. Development and Evaluation of the Anterior Cruciate Ligament Injury Score (ACLIS). *Orthopaedics & Traumatology*, Surgery & Research, 108(3), 103257. 10.1016/j.otsr.2022.103257

Maher, N. J., Brogden, C., Redmond, A. C., Siddle, H. J., Jones, G., Buck, D., Broadbent, S., Liversidge, G., Murr, J., Tingle, C., & Lunn, D. E. (2025). Disparity in anterior cruciate ligament injury management: a case series review across six National Health Service trusts. BMC Musculoskeletal Disorders, 26(1), 363. 10.1186/s12891-025-08572-5

McGrath, C., Palmgren, P. J., & Liljadahl, M. (2019). Twelve tips for conducting qualitative research interviews. Medical Teacher, 41(9), 1002–1006.

Nordenvall, R., Bahmanyar, S., Adami, J., Stenros, C., Wredmark, T., & Fellander-Tsai, L. (2012). A population-based nationwide study of cruciate ligament injury in Sweden, 2001-2009: incidence, treatment, and sex differences. American Journal of Sports Medicine, 40(8), 1808–1813. 10.1177/0363546512449306

Parwaiz, H., Teo, A. Q., & Servant, C. (2016). Anterior cruciate ligament injury: A persistently difficult diagnosis. Knee, 23(1), 116–120. 10.1016/j.knee.2015.09.016

Perera, N. S., Joel, J., & Bunola, J. A. (2013). Anterior cruciate ligament rupture: Delay to diagnosis. Injury, 44(12), 1862–1865. 10.1016/j.injury.2013.07.024

Robling, M. R., Pill, R. M., Hood, K., & Butler, C. C. (2009). Time to talk? Patient experiences of waiting for clinical management of knee injuries. Qual Saf Health Care, 18(2), 141–146. 10.1136/qshc.2007.022475

Sanders, T. L., Maradit Kremers, H., Bryan, A. J., Larson, D. R., Dahm, D. L., Levy, B. A., Stuart, M. J., & Krych, A. J. (2016). Incidence of Anterior Cruciate Ligament Tears and Reconstruction: A 21-Year Population-Based Study. American Journal of Sports Medicine, 44(6), 1502–1507. 10.1177/0363546516629944

Scott, S. M., Perry, M. A., & Sole, G. (2018). “Not always a straight path”: patients’ perspectives following anterior cruciate ligament rupture and reconstruction. Disability and Rehabilitation, 40(19), 2311–2317. 10.1080/09638288.2017.1335803

National Health Service. (2026). Knee pain https://www.nhs.uk/symptoms/knee-pain/

Simonsen, O., Jensen, J., Mouritsen, P., & Lauritzen, J. (1984). The accuracy of clinical examination of the knee joint. Injury, 16(2), 96–101.

Snoeker, B. A. M., Bakker, E. W. P., Kegel, C. A. T., & Lucas, C. (2013). Risk Factors for Meniscal Tears: A Systematic Review Including Meta-analysis. Journal of Orthopaedic and Sports Physical Therapy, 43(6), 352–367. http://search.ebscohost.com/login.aspx?direct=true&AuthType=ip,shib&db=sph&AN=88125925&site=ehost-live

Stutchbury, K. (2022). Critical realism: an explanatory framework for small-scale qualitative studies or an ‘unhelpful edifice’? International Journal of Research & Methods in Education, 45(2), 113–128.

Tong, A., Sainsbury, P., & Craig, J. (2007). Consolidated criteria for reporting qualitative research (COREQ): a 32-item checklist for interviews and focus groups. International Journal for Qualitative in Health Care, 19, 349–357.

von Essen, C., McCallum, S., Barenius, B., & Eriksson, K. (2020). Acute reconstruction results in less sick-leave days and as such fewer indirect costs to the individual and society compared to delayed reconstruction for ACL injuries. Knee Surgery, Sports Traumatology, Arthroscopy, 28(7), 2044–2052. 10.1007/s00167-019-05397-3

Watkins, R., Young, G., Western, M., Stokes, K., & McKay, C. (2020). Nobody says to you “come back in six months and we’ll see how you’re doing”: a qualitative interview study exploring young adults’ experiences of sport-related knee injury. BMC Musculoskeletal Disorders, 21(1), 419. 10.1186/s12891-020-03428-6

Whittaker, J. L., Chan, M., Pan, B., Hassan, I., Defreitas, T., Hui, C., Macedo, L., & Otto, D. (2020). Towards improving the identification of anterior cruciate ligament tears in primary point-of-care settings. BMC Musculoskeletal Disorders, 21(1), 252. 10.1186/s12891-020-03237-x

