## Supplementary material for "Patients’ Perspectives on the Contributors to and Impact of Delayed ACL Injury Diagnosis: A Qualitative Study": COREQ checklist

Supplementary File 1: COREQ checklist

Consolidated criteria for reporting qualitative studies (COREQ): 32-item checklist

Developed from:

Tong A, Sainsbury P, Craig J. Consolidated criteria for reporting qualitative research (COREQ): a 32-item checklist for interviews and focus groups. International Journal for Quality in Health Care. 2007. Volume 19, Number 6: pp. 349 – 357

| **Item No** | | **Guide Questions/Description** | **Reported on Page #** |  |  |
| --- | --- | --- | --- | --- | --- |
| **Domain 1: Research team and reflexivity** | | | |  |  |
| **Personal Characteristics** | | | |  |  |
| 1. Interviewer/ facilitator | | Which author/s conducted the interview or focus group? | Page 7 |  |  |
| 2. Credentials | | What were the researcher’s credentials? E.g., PhD, MD | Page 7 and 8 |  |  |
| 3. Occupation | | What was their occupation at the time of the study? | Page 7 and 8 |  |  |
| 4. Gender | | Was the researcher male or female? | Page 8 |  |  |
| 5. Experience and training | | What experience or training did the researcher have? | Page 7 and 8 |  |  |
| **Relationship with participants** | | | |  |  |
| 6. Relationship established | | Was a relationship established prior to study commencement? | Page 8 |  |  |
| 7. Participant knowledge of the interviewer | | What did the participants know about the researcher? e.g. personal goals, reasons for doing the research? | Page 9 |  |  |
| 8. Interviewer characteristics | | What characteristics were reported about the interviewer/facilitator? e.g. Bias, assumptions, reasons and interests in the research topic | Page 7 |  |  |
|  | **Domain 2: study design** | | |  |  |
|  | **Theoretical framework** | | |  |  |
|  | 9. Methodological orientation and Theory | What methodological orientation was stated to underpin the study? e.g. grounded theory, discourse analysis, ethnography, phenomenology, content analysis | Page 7 |  |  |
|  | **Participant selection** | | |  |  |
|  | 10. Sampling | How were participants selected? e.g., purposive, convenience, consecutive, snowball | Page 8 |  |  |
|  | 11. Method of approach | How were participants approached? e.g., face-to-face, telephone, mail, email | Page 8 |  |  |
|  | 12. Sample size | How many participants were in the study? | Page 10 |  |  |
|  | 13. Non-participation Setting | How many people refused to participate or dropped out? Reasons? | Page 10 |  |  |
|  | 14. Setting of data collection | Where was the data collected? e.g., home, clinic, workplace | Page 9 |  |  |
|  | 15. Presence of nonparticipants | Was anyone else present besides the participants and researchers? | Page 9 |  |  |
|  | 16. Description of sample | What are the important characteristics of the sample? e.g. demographic data, date | Page 10 |  |  |
|  | **Data collection** | | |  | No |
|  | 17. Interview guide | Were questions, prompts, and guides provided by the authors? Was it pilot tested? | Page 9 |  |  |
|  | 18. Repeat interviews | Were repeat interviews carried out? If yes, how many? | N/A – Page 9 indicates single interviews. |  |  |
|  | 19. Audio/visual recording | Did the research use audio or visual recording to collect the data? | Page 9 |  |  |
|  | 20. Field notes | Were field notes made during and/or after the interview or focus group? | Page 8 |  |  |
|  | 21. Duration | What was the duration of the interviews or focus group? | Page 10 |  |  |
|  | 22. Data saturation | Was data saturation discussed? | No, data sufficiency was noted in the Methods. |  |  |
|  | 23. Transcripts returned | Were transcripts returned to participants for comment and/or correction? | Page 10 |  |  |
|  | **Domain 3: analysis and findings** | | |  |  |
|  | **Data analysis** | | |  |  |
|  | 24. Number of data coders | How many data coders coded the data? | Page 7-8 |  |  |
|  | 25. Description of the coding tree | Did the authors provide a description of the coding tree? | Page 10 |  |  |
|  | 26. Derivation of themes | Were themes identified in advance or derived from the data? | Page 10 |  |  |
|  | 27. Software | What software, if applicable, was used to manage the data? | Page 9 |  |  |
|  | 28. Participant checking | Did participants provide feedback on the findings? | No |  |  |
|  | **Reporting** | | |  |  |
|  | 29. Quotations presented | Were participant quotations presented to illustrate the themes/findings? Was each quotation identified? e.g., participant number | Page 11-18 |  |  |
|  | 30. Data and findings consistent | Was there consistency between the data presented and the findings? | Page 11-18 |  |  |
|  | 31. Clarity of major themes | Were major themes clearly presented in the findings? | Page 11-18 |  |  |
|  | 32. Clarity of minor themes | Is there a description of diverse cases or a discussion of minor themes? | Page 11-18 |  |  |
