## Supplementary material for "Patients’ Perspectives on the Contributors to and Impact of Delayed ACL Injury Diagnosis: A Qualitative Study": Interview schedule

Supplement 2

PROPOSED INTERVIEW QUESTIONS TO PARTICIPANTS FOR THE SEMI- STRUCTURED INTERVIEWS

Participant: Alphanumeric ID.

Sex:

Age:

INTRODUCTION

Thank you for your time and for agreeing to take part, I appreciate this very much. As outlined in the participant information sheet we know that the ACL diagnosis has an average delay of 3-22 months. I aim to gather patient experience to help us identify the factors which lead to a delay in the diagnosis and to understand the impact of the delayed diagnosis on the person.

The following questions are designed to gather insight into your perspective and experience, which would help clinicians in the future to gain a deeper understanding on the issues, hence please be explicit and elaborate with your answers.

PERI INJURY

1. How did you injure your knee & what were your initial thoughts about your injury?

Prompts:

a. were you aware of anterior cruciate ligament injury at this point?

2. How long after the initial injury did you first present yourself to a healthcare practitioner?

Prompts:

a. What influenced the time between the injury and you visiting a healthcare practitioner.

3. What influenced your decision to present to the healthcare practitioner?

Prompts:

a. Discuss your symptoms and your understanding of them
b. Any suggestions from friends or professionals.
c. The information and knowledge you had about knee injuries
d. Any previous experience with knee injuries on yourselves or friends.

4. What setting did you first present to a healthcare professional? What influenced this choice?

Prompts:

a. Was it a GP, nurse practitioner, physiotherapist, Emergency department doctor?
b. Why did you decide to present to this professional?

HEALTHCARE

5. How did you feel your appointment went during the initial consultation?

Prompts:

a. Were your concerns addressed?
b. Was there any aspect of this appointment that you felt positive about?
c. Were there any aspects of this appointment that you felt negative about?

6. Did you feel confident with the healthcare professional’s decision and advice?
   a. Can you explain on what factors influenced on you being satisfied or not satisfied with the decision on your diagnosis and management?
   b. Did you feel the professional was competent and informed on what he/ she was dealing with?
7. What impact did the initial consultation have on your future decisions on presenting yourselves to further examination of your injured knee?

Prompts:

a. Did any of the experience from your initial consultation encourage or discourage you from presenting for further examination?

8. At what stage after the injury did you get a confirmed diagnosis of ACL injury and how many healthcare professionals had you seen by then?
9. If you had seen more than one healthcare professional to reach a suspected or a confirmed ACL injury what factors influenced, you to gain further or repeated advice from healthcare professionals?

Prompt:

a. Please comment on physical symptoms.
b. Please comment on emotional consequences.
c. Do you think any aspect of your care could have facilitated a quicker diagnosis?

IMPACT OF THE INJURY ON THE RESEARCH PARTICIPANT

10. How did the delayed diagnosis of the injury affect you? Prompts:
    a. Please comment on sports, work, family life, emotional impact and any other aspects.
11. What aspects of your overall experience do you think could have facilitated a quicker diagnosis?

Prompts:

a. Please think of things you could have known or done better and what others could have known or done better.

12. What changes in your experience with the injury do you think could have reduced the impact of the injury on you?

Prompts:

a. Please comment on sports, work, family life, emotional impact and any other aspects.

13. Is there anything else you would like to add?
